# Oral-Cardiovascular Links via Blood Biomarkers: An Exposure-Wide Analysis

**DOI:** 10.64898/2026.03.13.26348364

**Authors:** Yanbei Lu, Zhiheng Yi, Yanjia Zhuang, Dasen Yuan, Lei Lei, He Cai, Tao Hu

## Abstract

**Objectives:** Cardiovascular diseases (CVDs) remain a global health priority, with oral health linked to cardiovascular risk. However, existing studies have mainly focused on limited periodontal measures and lack analyses of multidimensional oral health.

**Methods:** Using data from 4,564 participants in the China Multi-Ethnic Cohort, this study employed an exposure-wide analysis to systematically examine associations between multidimensional oral health indicators and multiple CVD-related outcomes, and to evaluate the role of blood and urine-based biomarkers as potential mediators.

**Results:** We found that indicators regarding caries, periodontal status, and occlusal function were significantly associated with carotid plaque prevalence. Sex-stratified analyses confirmed that all significant associations were concentrated in females, instead of males. Age-stratified analyses showed distinct patterns, with meaningful associations concentrated among participants under 65, where indicators regarding caries, periodontal status, and occlusal function were related to the FRS. In adults aged 65 and older, only the number of functional tooth units was associated with carotid plaque. Mediation analyses identified multiple blood biomarkers, including hemoglobin A1c and Platelet Large Cell Ratio, as partial mediators.

**Conclusions:** These findings indicate that multidimensional oral health correlates with cardiovascular outcomes, with obvious sex and age differences, and that blood biomarkers play an important mediating role. Our findings provide new insights that extend beyond traditional periodontal associations by highlighting the relevance of multidimensional oral health to cardiovascular risk, supporting its consideration in cardiovascular prevention.

## 1. Introduction

Oral health, a crucial component of overall health, is increasingly recognized as interacting with systemic diseases, and cardiovascular diseases (CVDs) persist as a dominant global public health threat closely linked to oral health status ^1^. Among middle-aged and older adults, CVDs affect 40%–60% of individuals, coexisting with atherosclerosis, hypertension, and coronary artery disease. Moreover, the unfavorable trends in cardiometabolic risk factors are no longer confined to this population. Alarmingly, a pattern of deteriorating cardiovascular health, marked by rising rates of obesity and diabetes alongside worsening hypertension control, is now distinctly observable among younger individuals ^1^. This phenomenon is also associated with poor oral health and results in high disability and socioeconomic burdens. As a key non-traditional CVD risk factor, oral health interacts with cardiovascular conditions, emphasizing the need to clarify their interconnections ^2,3^.

Notably, European CVD prevention guidelines have classified periodontitis as a potential risk-enhancing factor for CVD ^4,5^. This linkage is largely driven by chronic systemic inflammation and ectopic oral microbiota colonization, which activate inflammation, damage vascular endothelial cells, and promote oxidative stress ^6^. Aside from periodontitis, a large population-based cohort found that dental caries and tooth loss remained significantly associated with major adverse cardiovascular events after adjustment for co-morbidity factors (MACE) ^3^. As a key CVD pathogenic event, thrombosis can be induced by Streptococcus mutans, which alters platelet activation, aggregation, and coagulation in endothelial cells and increases neutrophil migration ^7^. Additionally, NHANES III analyses show that favorable cardiovascular health reduces all-cause mortality only in denture-wearing edentulous older adults, an association that may be partly attributable to improved dietary patterns and nutrient intake from better masticatory function ^8,9^. However, existing studies have rarely examined the impact of CVD across different oral conditions simultaneously, overlooking the multidimensional and coexisting nature of oral diseases.

This study aimed to use an exposure-wide approach to systematically investigate associations between oral health as a non-traditional cardiovascular risk factor and cardiovascular disease. We further examined whether these associations varied across age and sex groups, and evaluated the mediating role of blood biomarkers by assessing metabolic indicators, cardiac injury biomarkers, and systemic inflammatory markers in the relationships between oral health and CVD outcomes. This study provides population-based evidence to support the incorporation of comprehensive oral health indicators into cardiovascular risk assessment beyond traditional risk factors.

## 2. Methods

The source data supporting the findings of this study have been deposited in the Zenodo, and are available at: https://zenodo.org/records/18976861. Due to ongoing related research, the data are under embargo and will be made publicly available in March 2031. During the embargo period, the data can be made available from the corresponding author upon reasonable request.

### 2.1 Study Design and Data Collection

Data regarding questionnaires, oral examination, physical examination, laboratory examination, and carotid ultrasound were derived from the 2024 follow-up (Clinical trial registration number: ChiCTR2400082963) of the China Multi-Ethnic Cohort (CMEC), a population-based cohort established in 2018 across five southwestern Chinese provinces, spanning an altitude range from 400 to 3650 meters ^10^. The primary objective of the current study was to examine whether oral health may influence CVD risk in this ethnically and altitudinally diverse population. In accordance with epidemiological reporting standards, this study strictly adhered to the Strengthening the Reporting of Observational Studies in Epidemiology (STROBE) guidelines throughout all study phases, including design implementation, data harmonization, and analytical validation. The privacy rights of human subjects have been observed.

Questionnaire data were collected through face-to-face interviews by trained researchers using electronic questionnaires. Oral examination was conducted by experienced and trained dentists via chairside examination. Local community hospitals collected physical examination and ultrasound data following standardized training and instrument calibration. Laboratory examination data were gathered by a third-party company with appropriate national qualifications. All procedures were performed in compliance with relevant laws and institutional guidelines and have been approved by the appropriate institutional committee(s). This study adhered to the Declaration of Helsinki, and all participants signed informed consent forms prior to data collection. Ethical approval was obtained from the Medical Ethics Review Committee of Sichuan University (WCHSIRB-D-2023-225-R1).

### 2.2 Eligibility Criteria

Individuals who completed the 2024 CMEC follow-up survey and had complete and valid oral examination data were eligible for inclusion in the current study. Participants were further required to have available cardiovascular-related data, including variables required for calculating the FRS, ultrasound-confirmed carotid plaque assessment, and hypertension information. All participants should meet the original CMEC eligibility criteria, including being permanent residents aged 30–79 years at baseline (18–79 years for Tibetans) who were capable of completing study procedures and participated in standardized questionnaire interviews, physical examinations, and blood tests during the cohort surveys.

Participants were excluded if they had incomplete follow-up data in 2024, missing or invalid oral examination data, met the original CMEC exclusion criteria (e.g., severe physical or mental illness or non-compliance with study requirements), withdrew informed consent or were lost to follow-up before the oral examination.

### 2.3 Variables

*Exposures* The oral exposure factors included multiple aspects of dental caries, periodontal status, oral prosthesis, oral function, oral frailty, and xerostomia. Dental status was evaluated through chairside clinical examination, covering coronal and root caries as well as fillings, with the DMFT index calculated to assess overall caries experience; residual roots (teeth with most or all clinical crown missing) were also counted. Periodontal status was examined using a CPI probe, including evaluations of periodontal pockets, deep periodontal pockets, bleeding on probing, and periodontitis (including severe periodontitis) based on probing depth and clinical attachment loss (AL). Additionally, other factors assessed included the number of tooth loss, functional dentition, the number of functional tooth loss, occlusal pairs, functional tooth units (FTU), dental prosthesis use (e.g., crowns, implants, dentures), swallowing function, and oral frailty (assessed via the OFI-8 questionnaire with a score ≥ 4 indicating oral frailty).

#### Outcomes

The outcome measures included the Framingham Risk Score (FRS), carotid plaque, and hypertension. FRS was calculated based on age, high-density lipoprotein cholesterol, total cholesterol, systolic blood pressure, smoking status, and diabetes status, with an FRS of ≥ 10% defined as elevated cardiovascular risk ^11^. Carotid plaque was determined by the presence of plaque reported in cervical vascular ultrasound examination results. Hypertension was defined as an average systolic blood pressure ≥ 140 mmHg or an average diastolic blood pressure ≥ 100 mmHg.

#### Confounders

The confounding factors included demographic and socioeconomic factors, Body Mass Index (BMI), duration of sedentary behavior, smoking status, and diabetes. Demographic and socioeconomic factors (including age, sex, education level, and income level) were collected via questionnaire. BMI was calculated based on physical measurements. Weekly sedentary time was also obtained from questionnaires. Current smoking was defined as having smoked a total of more than 100 cigarettes and currently smoking (excluding those who had quit for more than six months). Diabetes was defined as having a glycated hemoglobin (HbA1c) level ≥ 6.5% or current use of anti-diabetic medication.

#### Mediators

The mediators were derived from fasting venous blood samples and urine samples analyzed via standardized laboratory assays, which provided data for the listed hematological, biochemical, inflammatory, myocardial-related tests and urine tests. This study established standardized procedures during the data collection phase to ensure technical consistency among operators through structured training and implemented the calibration mechanism for equipment to minimize systematic errors.

### 2.4 Statistics

Statistical analysis was performed using Python 3.11.0 software. For descriptive statistics, continuous variables were reported based on normality (assessed via the D’Agostino test and skewness-kurtosis criteria), normally distributed data as mean ± standard deviation, and non-normally distributed data as median (interquartile range). Categorical variables were presented as counts (percentages). The data were original and had not undergone any modifications such as imputation. Except for the baseline data, multiple imputation was performed for covariates and mediating variables in all other data included in the analysis. The odds ratios (ORs) for cardiovascular outcomes were estimated using multivariable logistic regression. To address potential multicollinearity, multivariable logistic regression models incorporated Benjamini-Hochberg false discovery rate (FDR) correction. A stepwise covariate adjustment approach was adopted to account for potential confounding factors across sequentially constructed models. A comprehensive set of confounders was gradually incorporated, including demographic characteristics, socioeconomic status indicators, lifestyle factors, and cardiometabolic comorbidities. Hypertension was additionally adjusted for when the outcome was the FRS or carotid plaque. For stratified analyses, logistic regression models were stratified by sex (male/female) and age (< 65 years/≥ 65 years), with the stratifying variable excluded from covariates in respective subgroup models.

Mediation analyses were performed only for exposure-outcome pairs with statistically significant total effects. Bootstrap mediation analysis quantified direct, indirect, and total effects of oral health metrics on cardiovascular outcomes via biochemical mediators (e.g., Platelet Large Cell Ratio (P-LCR), HbA1C). For statistical criteria, all tests were two-sided; *P* < 0.05 indicated statistical significance, and *P* < 0.05 (after FDR correction) defined significant associations.

## 3. Results

### 3.1 Characteristics of Participants

In total, 4564 participants were included in this study, with an average age of 58.00±10.74 years. Among them, 38.91% were male, 61.09% were female. Regarding cardiovascular-related indicators, 41.56% had hypertension, 15.16% showed carotid plaques on ultrasonography, and the average FRS was 0.13±0.09 (Fig. 1, Table 1, and Table S1).

**Fig. 1:**
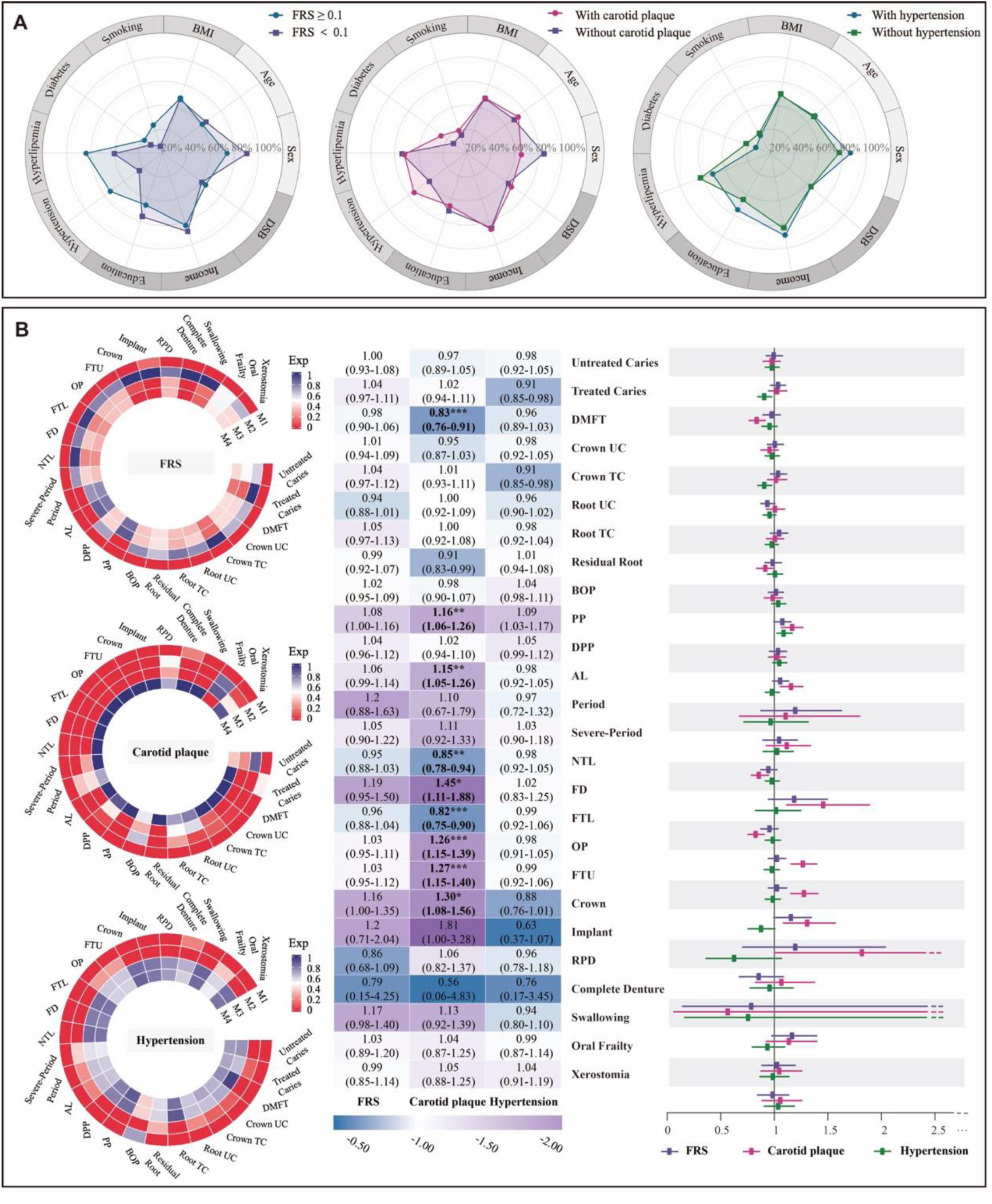
Associations Between Oral Health Indicators and Cardiovascular Outcomes (FDR-Corrected Logistic Regression). (A) Radar charts illustrating demographic/clinical covariate distributions (age, sex, body mass index, smoking, diabetes, hyperlipidemia, hypertension, education, income, duration of sedentary behavior) across three cardiovascular risk strata. Radial axes represent covariate prevalence. (B) Left: integrated FDR-corrected regression results, three circular heatmaps display FDR-significant P-values for oral health exposures across Models 1–4 (Model 1: unadjusted; Model 2: adjusted for age and sex; Model 3: further adjusted for body mass index, smoking, diabetes, and hyperlipidemia, with hypertension additionally included only when outcomes were FRS or carotid plaque; Model 4: additionally adjusted for education, income, and duration of sedentary behavior); Middle: presents odds ratios (ORs) and 95% confidence intervals (CIs); Right: forest plot visualizes ORs and 95% Cis. \**P* < .05, \*\**P* < .01, \*\*\**P* < .001 (FDR-adjusted *P*). *AL* Attachment loss, *BOP* Bleeding on probing, *DSB* Duration of sedentary behavior, *DPP* Deep periodontal pocket, *FD* Functional dentition, *FTL* Functional tooth loss, *NTL* Number of tooth loss, *OP* Occlusal pairs, *FTU* Functional tooth units, *Period* Periodontitis, *PP* Periodontal pocket, *Severe*-*Period* Severe periodontitis, *TC* Treated caries, *UC* Untreated caries.

**Table 1.**
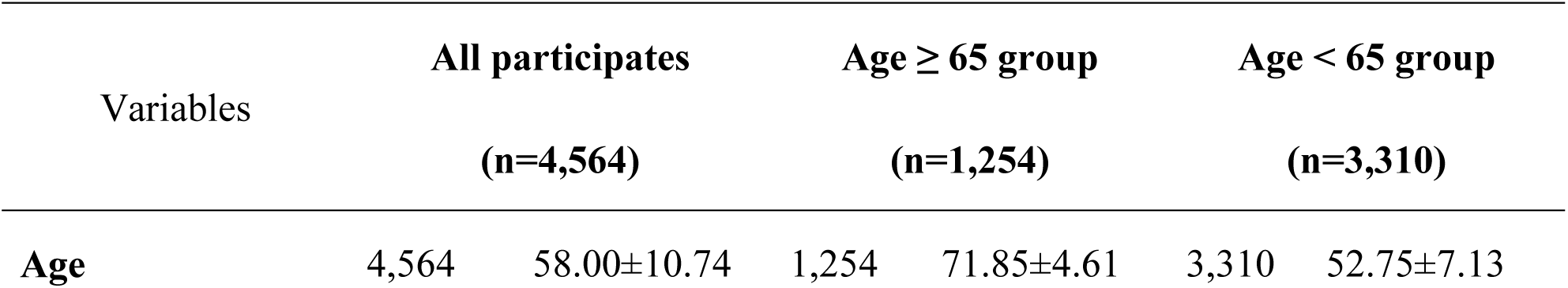
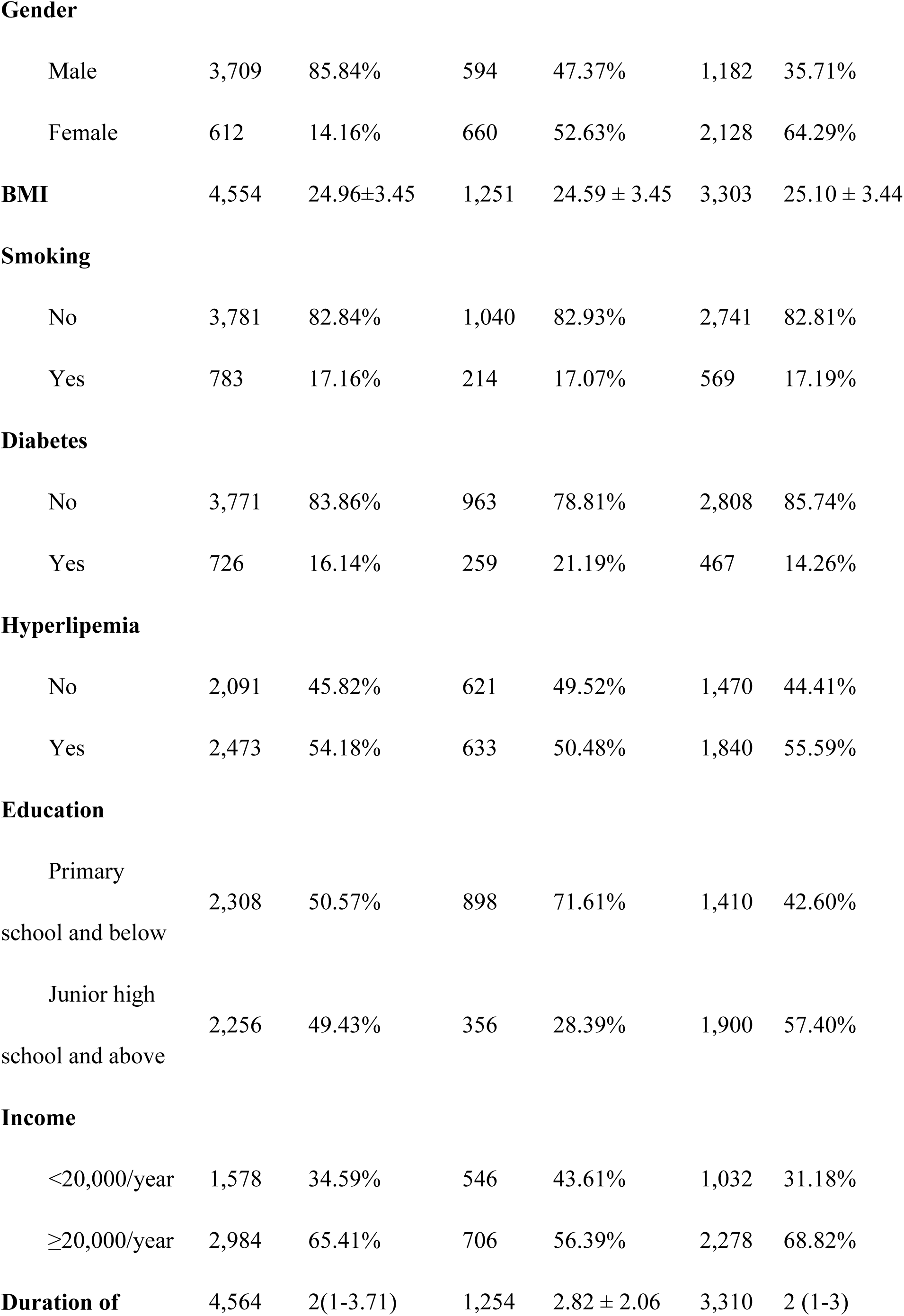
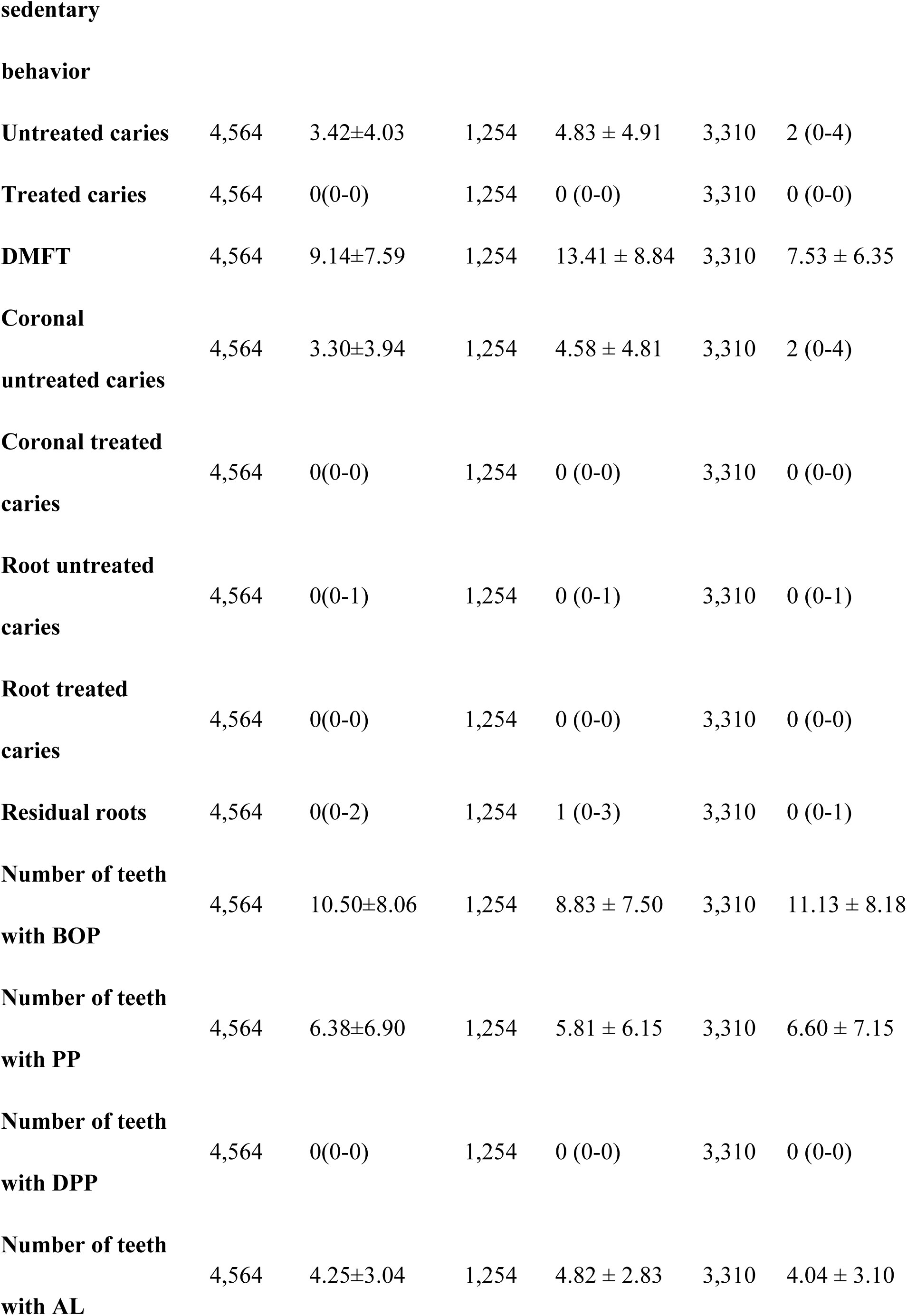
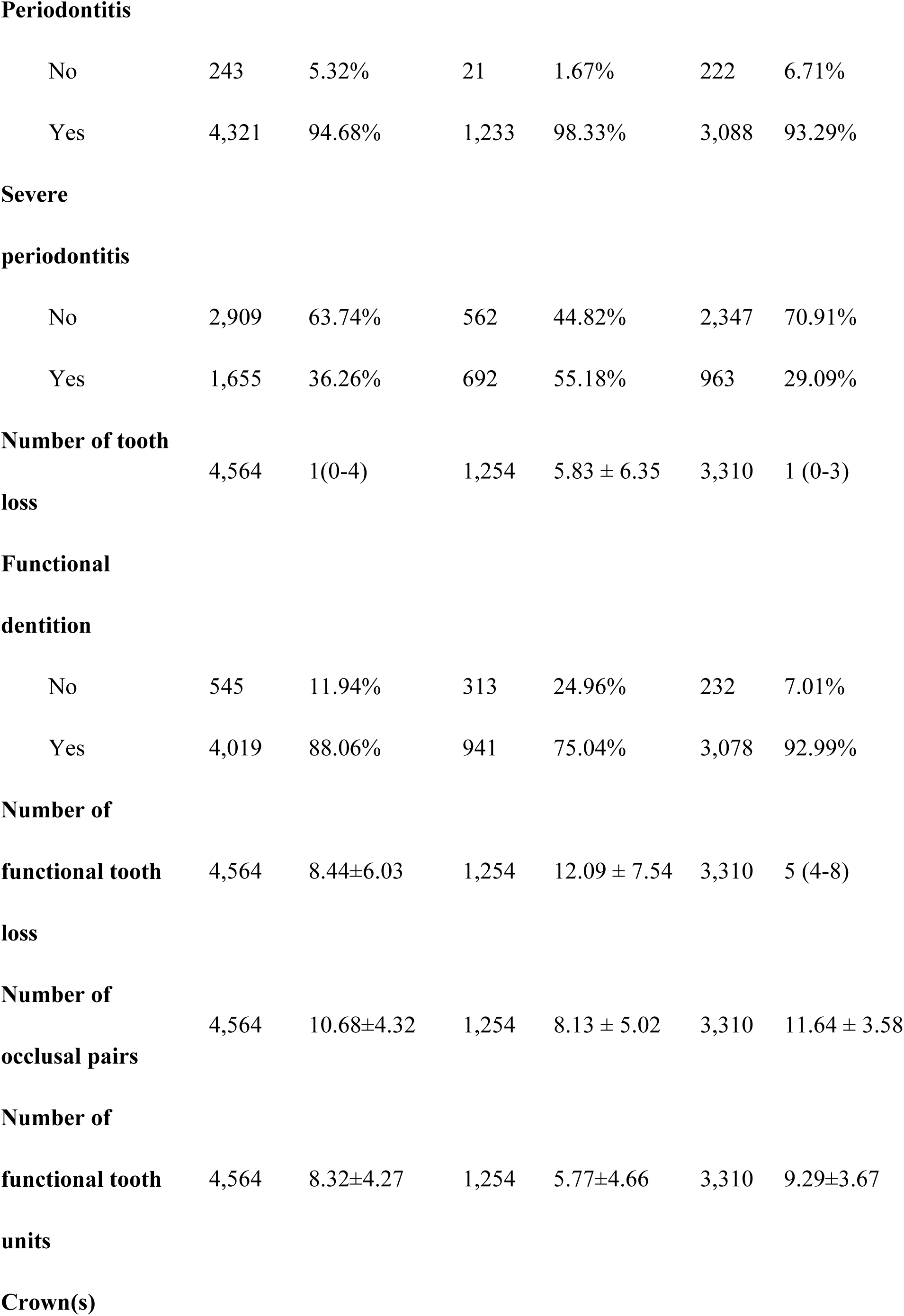
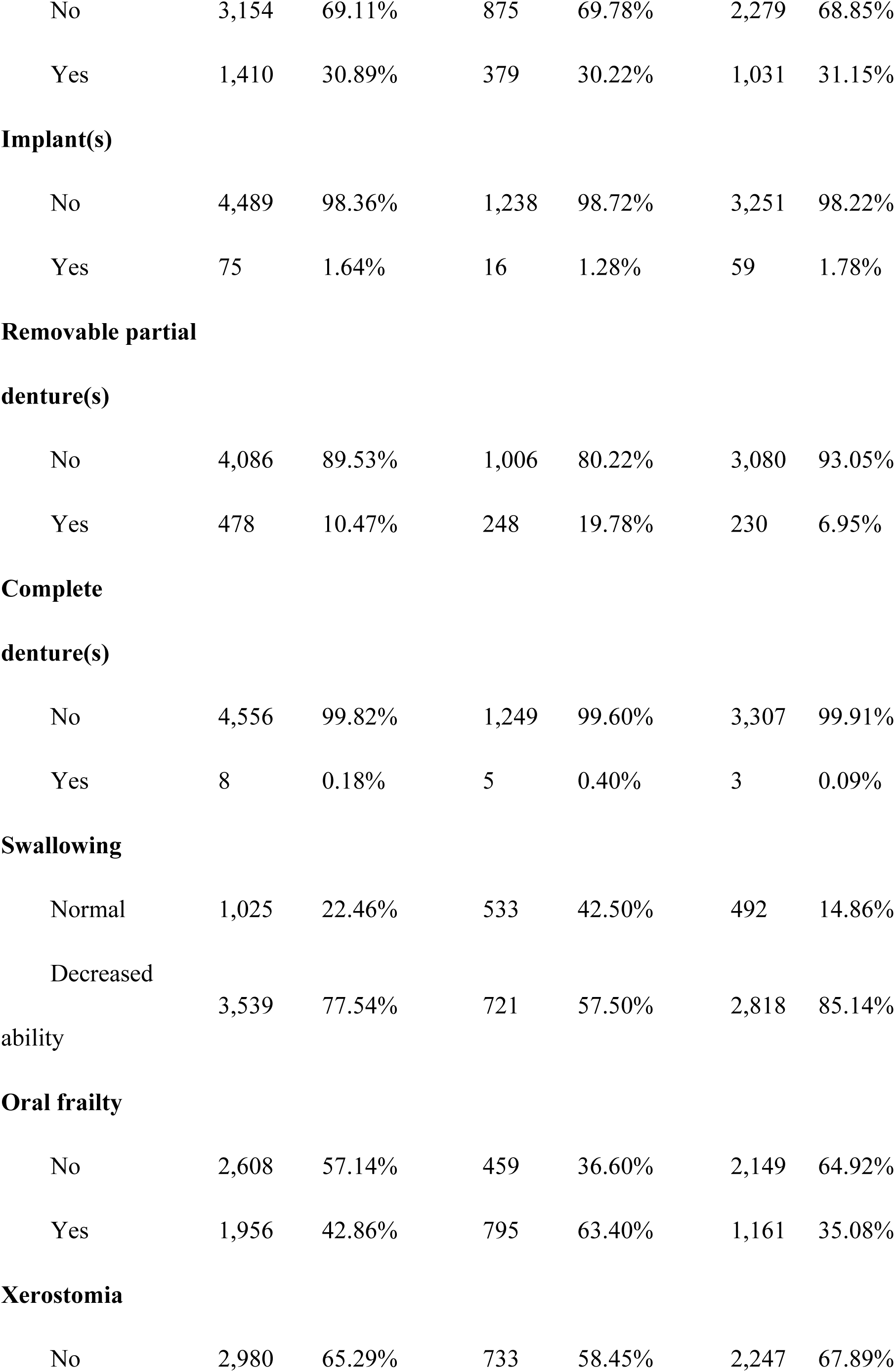
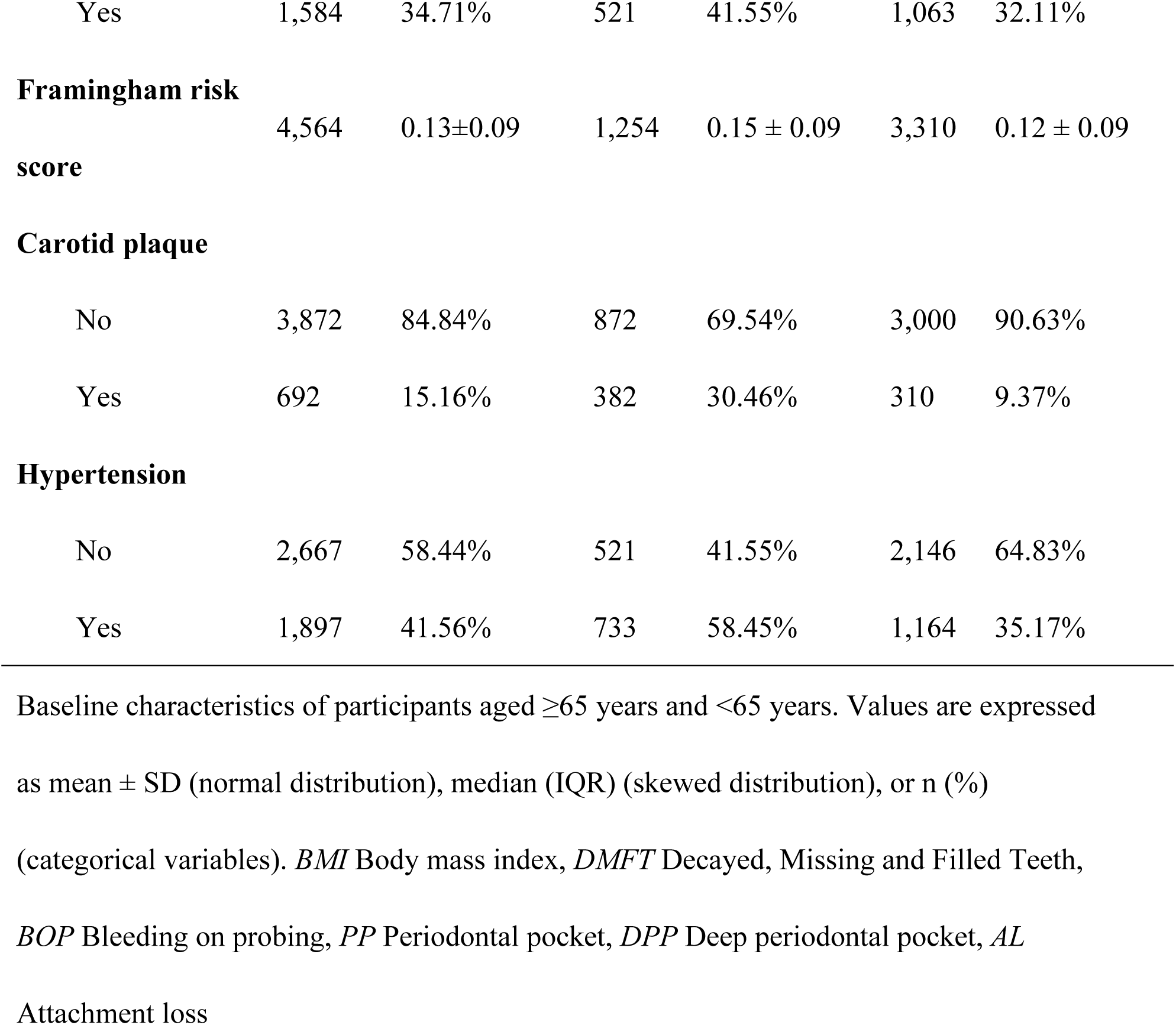
Characteristics of the study participants (n=4,564)

### 3.2 Associations between oral health and cardiovascular-related indicators

After sequential adjustment for covariates (age, sex, BMI, smoking, diabetes, hyperlipidemia, hypertension, education, income, and duration of sedentary behavior) and FDR correction, several oral health indicators were found to be significantly associated with the presence of carotid plaque. Specifically, caries-related indicators (DMFT: OR 0.83, 95%CI 0.76 to 0.91) showed a negative association with the presence of carotid plaque. In contrast, a more severe periodontal status (greater number of teeth with PP: 1.16, 1.06 to 1.26; greater number of teeth with AL: 1.15, 1.05 to 1.26) was positively associated with the presence of carotid plaque. For indicators reflecting occlusal function, better preserved occlusal function (functional dentition: 1.45, 1.11 to 1.88; more occlusal pairs: 1.26, 1.15 to 1.39; greater number of FTU: 1.27, 1.15 to 1.40; use of crowns: 1.30, 1.08 to 1.56) was associated with a higher prevalence of carotid plaque, and vice versa (greater tooth loss: 0.85, 0.78 to 0.94; greater functional tooth loss: 0.82, 0.75 to 0.90; Fig. 1 and Table S2).

However, in the overall population, no statistically significant associations were observed between oral health indicators (including caries status, periodontal status, occlusal function, oral prostheses, swallowing function, oral frailty, and xerostomia) and hypertension or FRS.

### 3.3 Associations Stratified by Sex and Age

The sex-stratified analysis revealed that all statistically significant associations were concentrated in the female population. For the outcome of carotid plaque, caries-related indicators (DMFT: OR 0.76, 95% CI 0.66 to 0.87) had a negative association; for periodontal status, a greater number of teeth with PP (1.16, 1.03 to 1.31) was positively associated; and regarding occlusal function and tooth loss, greater tooth loss (0.75, 0.65 to 0.86) and greater functional tooth loss (0.73, 0.64 to 0.84) were negatively associated, while better preserved occlusal function (functional dentition: 1.72, 1.18 to 2.49; more occlusal pairs: 1.37, 1.19 to 1.57; greater number of FTU: 1.42, 1.24 to 1.64) was associated with a higher prevalence of carotid plaque. Consistent with the overall population, no significant associations were observed between the other two outcome indicators (hypertension and FRS) and oral health indicators in the female population (Fig. 2 and Table S3).

**Fig. 2:**
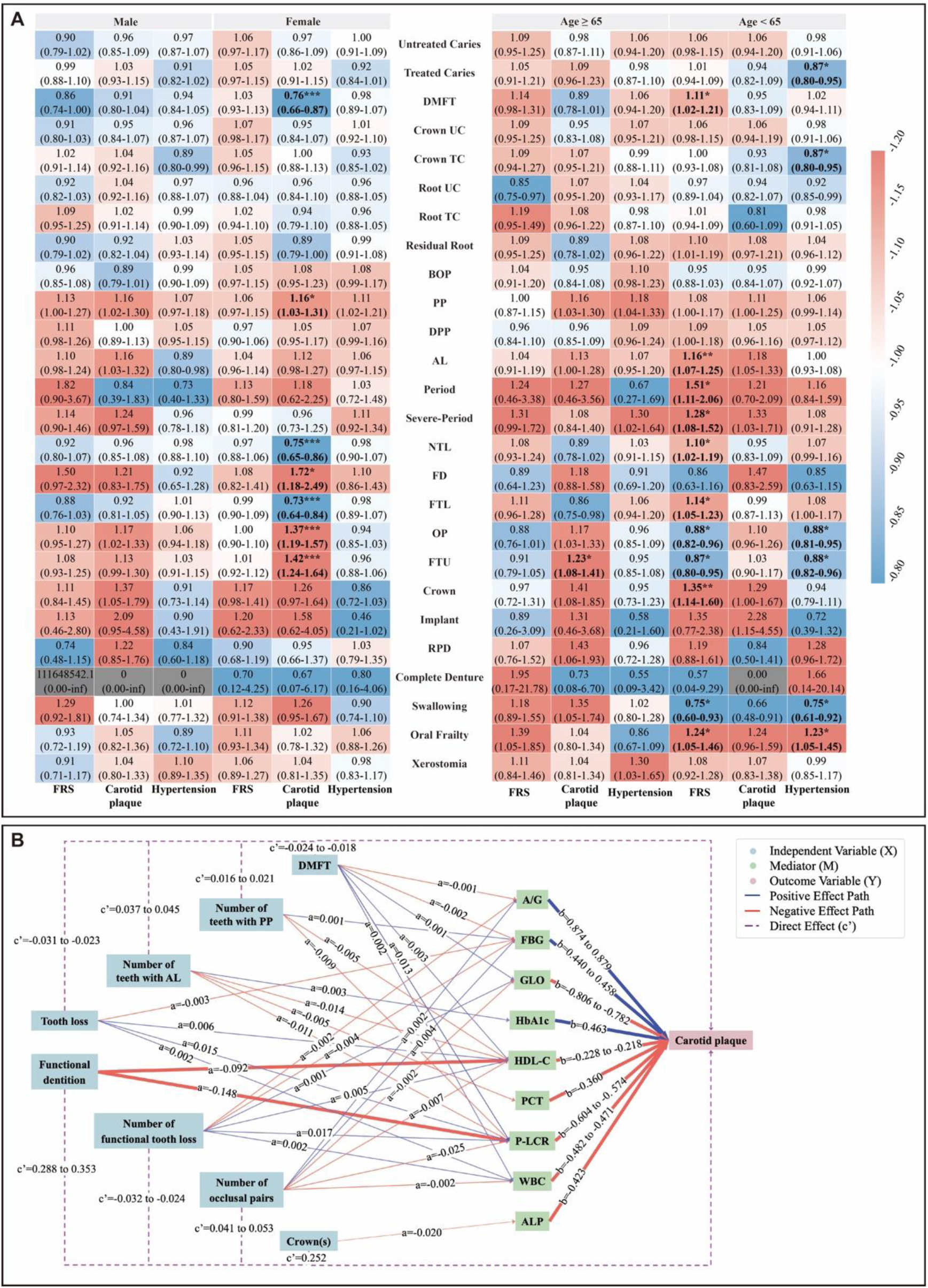
Results of stratified analysis and mediation analysis of oral health indicators in relation to cardiovascular-related outcomes. (A) Heatmaps of sex and age-stratified odds ratios (ORs) and 95% confidence intervals (CIs) for oral health indicators in relation to cardiovascular outcomes. (B) Mediation analysis of oral health indicators and cardiovascular outcomes via metabolic and inflammatory biomarkers. \**P* < .05, \*\**P* < .01, \*\*\**P* < .001 (FDR-adjusted *P*). *AL* Attachment loss, *BOP* Bleeding on probing, *DPP* Deep periodontal pocket, *FD* Functional dentition, *FTL* Functional tooth loss, *NTL* Number of tooth loss, *OP* Occlusal pairs, *OU* Occlusal units, *Period* Periodontitis, *PP* Periodontal pocket, *Severe*-*Period* Severe periodontitis, *TC* Treated caries, *UC* Untreated caries, *A/G* Albumin/Globulin ratio, *ALP* Alkaline Phosphatase, *FBG* Fasting Blood Glucose, *GLO* Globulin, *HbA1C* Hemoglobin A1C, *HDL-C* High-Density Lipoprotein Cholesterol, *P-LCR* Platelet Large Cell Ratio, *PCT* Procalcitonin, *WBC* White Blood Cell.

Age-stratified analysis revealed different association patterns between oral health indicators and cardiovascular-related outcomes for participants aged ≥65 years and <65 years. For participants aged ≥65 years, no significant associations were observed between oral health indicators and cardiovascular-related outcomes, except that a greater number of FTU (OR 1.23, 95% CI 1.08 to 1.41) showed a significant positive association with the presence of carotid plaque.

In contrast, multiple oral health indicators retained statistical significance in participants <65 years. For FRS, indicators reflecting poor oral health status, including higher DMFT (1.11, 1.02 to 1.21), a greater number of teeth with AL (1.16, 1.07 to 1.25), presence of periodontal condition (1.51, 1.11 to 2.06), severe periodontitis (1.28, 1.08 to 1.52), greater tooth loss (1.10, 1.02 to 1.19), greater functional tooth loss (1.14, 1.05 to 1.23), presence of crown(s) (1.35, 1.14 to 1.60), and oral frailty (1.24, 1.05 to 1.46) were positively associated with medium-to-high 10-year cardiovascular risk, whereas a greater number of occlusal pairs (0.88, 0.82 to 0.96), a greater number of FTU (0.87, 0.80 to 0.95), and better swallowing ability (0.75, 0.60 to 0.93) were negatively associated with this outcome. Regarding hypertension, more active treatment of oral caries was negatively associated with hypertension risk, including caries treatment (more filled teeth: 0.87, 0.80 to 0.95; more coronal fillings: 0.87, 0.80 to 0.95), better occlusal status (more occlusal pairs: 0.88, 0.81 to 0.95; greater number of FTU: 0.88, 0.82 to 0.96). Whereas oral frailty (1.23, 1.05 to 1.45) positively associated with hypertension risk. In addition, better swallowing ability was negatively associated with both carotid plaque (0.66, 0.48 to 0.91) and hypertension (0.75, 0.61 to 0.92; Fig. 2 and Table S4).

### 3.4 Mediation Analysis

Mediation analyses performed revealed that multiple biochemical indicators played significant partial mediating roles in the associations between oral health indicators and carotid plaque (Table 2). P-LCR showed relatively high mediation proportions in all related indicators, with 31.78% for DMFT, 19.30% to 41.38% for periodontal health indicators and 14.76% to 31.49% for occlusal function related indicators. For periodontal health indicators, High-Density Lipoprotein Cholesterol (HDL-C) (mediation proportion 5.48% to 6.99%) showed partial mediating effects on both periodontitis related indicators. Besides, Fasting Blood Glucose (FBG) (5.64%) had a partial mediating effect on Number of teeth with PP, Procalcitonin (PCT) (4.37%) and HbA1C did so on Number of teeth with AL. Regarding DMFT and occlusal function related indicators (Tooth loss, Functional dentition, Number of functional tooth loss, Number of occlusal pairs), inflammation/immune-related indicators (White Blood Cell count (WBC), Globulin (GLO), Albumin/Globulin ratio (A/G)) showed partial mediating effects with proportions ranging from 2.08% to 3.49%. Consistent with this, glucose metabolism indicators FBG (3.21% to 4.77%) and lipid metabolism indicator HDL-C (2.52% to 5.63%) also exhibited partial mediating effects. Additionally, for Crown(s), Alkaline Phosphatase (ALP; 3.67%) exhibited a significant partial mediating effect.

**Table 2.**
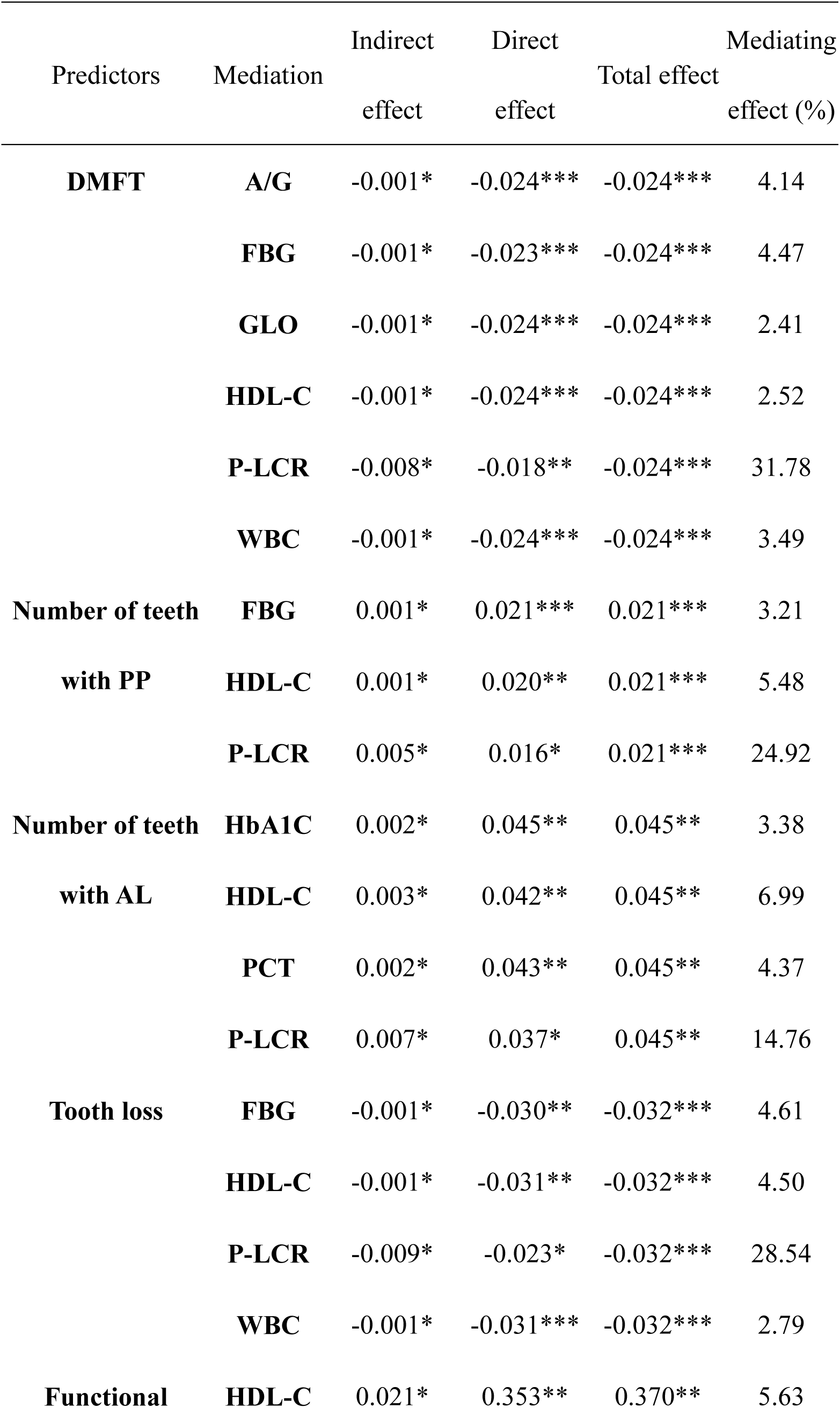
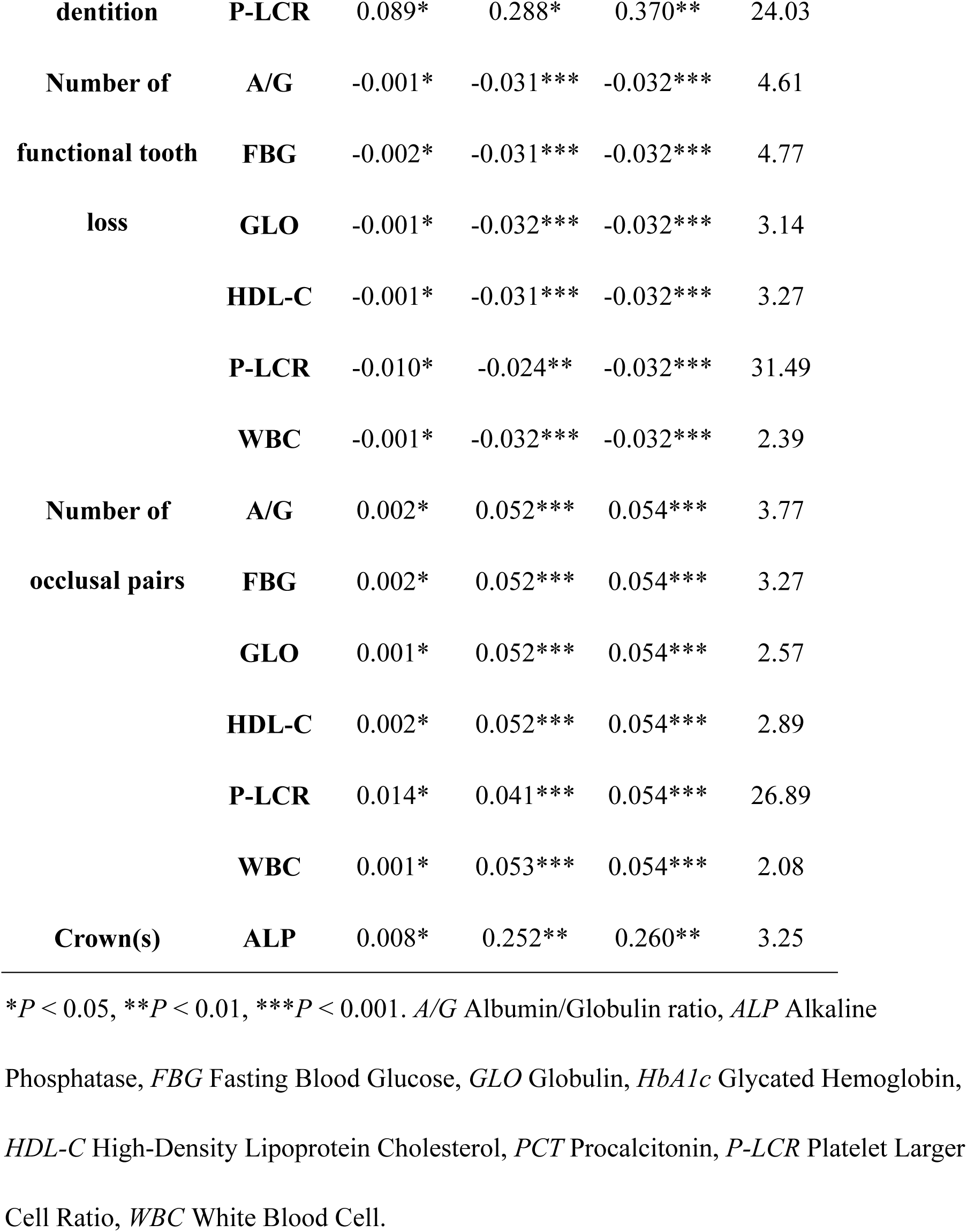
Biochemical indicators as partial mediators for oral health and carotid plaque.

## 4. Discussion

Multidimensional oral health, as a non-traditional cardiovascular risk factor domain, is closely linked to cardiovascular disease risk, with caries, periodontal status, occlusal function, and oral prosthesis emerging as significant factors across the total population, age, and gender stratifications. For the total population and female population, these associations mainly focus on carotid plaques, while in the population under 65 years old, they are mainly focused on long-term cardiovascular risk (FRS). Metabolic and inflammatory indicators partially explained these associations. These findings highlight that multidimensional oral health was closely related to cardiovascular risk, which differs in short-term and long-term cardiovascular risk among various sex and age groups.

Our analysis revealed significant correlations between DMFT, the severity of periodontitis, the status of occlusal function, and cardiovascular risk. This relationship persisted after strict adjustment for confounding factors, indicating that different types of oral diseases are measurable dimensions associated with the risk of cardiovascular diseases. This highlights the importance of oral health management in the prevention and prognosis of cardiovascular health. For the total population, better oral occlusal function and the use of crown restorations were positively correlated with carotid plaques, while DMFT was negatively correlated with carotid plaques. Adequate occlusal support contributes to efficient chewing, which may affect dietary quality (e.g., preference for the intake of pro-inflammatory substances such as red meat), nutrient absorption, and systemic stress responses ^12,13^. These factors may jointly regulate systemic inflammation and traditional cardiovascular risk characteristics, providing a reasonable nutritional and physiological basis for the observed associations ^14^. Periodontitis was associated with higher cardiovascular risk in all population stratifications, which is consistent with the mainstream view that oral conditions mediate chronic systemic inflammation and ectopic colonization of oral microbiota ^6^.

Sex- and age-stratified analyses revealed significant heterogeneity across various subgroups. Significant associations were predominantly in females, with effect directions consistent with the total population, suggesting greater susceptibility to poor oral health-induced systemic inflammation/metabolic disorders. Indeed, after the menopausal transition, the incidence of coronary heart disease (CHD) in women increases, suggesting a role for estrogen in the prevention of CVD ^15^. Regarding age stratification, oral health was associated with CVD risk primarily in individuals under 65 years. This pattern suggests that young adulthood and middle adulthood may represent a pivotal window during which the cumulative burden and functional sequelae of oral diseases may partly contribute to the pathogenesis of both subclinical and clinical cardiovascular disease ^16–18^. Conversely, age-related comorbidities, cumulative lifetime risk factors, and survival bias may mask such associations in those over 65 years. ^3^. Interestingly, the direction of associations between occlusal function/DMFT and cardiovascular diseases differed in individuals <65 years versus total/female populations. Beyond a certain occlusal decline threshold, fruit/vegetable (not processed food) intake may affect nutrient absorption/systemic stress ^12,13^.

For all significant combinations, the thrombosis-related indicator P-LCR exhibited the highest mediating proportion. FBG and HbA1c highlight the roles of glucose metabolic dysregulation in the oral-cardiovascular axis. Liver metabolism-related indicators (ALP) and bacterial infection indicator (PCT) reinforce the impact of persistent low-grade systemic inflammation. The mediating effects of WBC count, GLO, and A/G ratio (related to inflammation and immune response) further support this. Chronic oral inflammation and pathogenic plaques may induce systemic insulin resistance and glucose homeostasis disruption ^19^. Metabolic disorders combined with persistent low-grade systemic inflammation may impair vascular endothelial function, and increased oxidative stress and advanced glycation end products accelerate atherosclerosis and hypertension ^20^. As a component of Virchow’s triad, endothelial dysfunction directly activates arterial thrombosis and venous thromboembolism (VTE), which triggers plaque rupture in subsequent thrombosis ^21^.

Periodontitis patients exhibit elevated circulating thrombotic factor concentrations and increased blood cell reactivity, indicating that periodontitis may directly promote thrombotic disease progression and increase carotid plaque rupture risk ^6^. These findings suggest the oral health-CVD association is likely multiplex, concurrently mediated by metabolic dysregulation and inflammatory pathways.

Using data from the China Multi-Ethnic Cohort, this study provides an exposure-wide evaluation of oral health in relation to cardiovascular-related outcomes, highlighting caries, periodontitis, and occlusal function as potential non-traditional risk factors for cardiovascular risk stratification.

### Limitations

However, several limitations of this study should be acknowledged. Although potential confounders were adjusted for, the cross-sectional design fundamentally limits causal inference, and reverse causation remains possible. To further address this limitation, continued follow-up of the CMEC cohort is planned, and the next assessment within 3–5 years will provide an opportunity to prospectively examine the temporal sequence and direction of the observed associations. Another consideration is the reliance on clinically assessed oral health indicators, which, though systematic, might not capture all subclinical conditions or historical disease burden. These aspects will also be considered for inclusion in the next follow-up data collection to enable a more refined and longitudinal characterization of oral health trajectories. Finally, by assessing biomarker-based biological pathways, this study identified metabolic and inflammatory blood biomarkers as partial mediators of the association between oral health and cardiovascular disease risk. Future studies integrating both biomarkers and other plausible mediators, such as behavioral factors (e.g., oral hygiene practices and dietary patterns) and broader nutritional status, will be important for a more comprehensive understanding of the oral–cardiovascular connection.

## 5. Conclusion

This study provides evidence from an exposure-wide analysis that synthesizes multidimensional oral health indicators into a unified analytical dimension. It links oral health to cardiovascular disease (CVD) risk in a multi-ethnic Chinese population, with caries, periodontal status, and occlusal function emerging as the key findings. Sex- and Age-specific patterns, together with identified mediating pathways involving metabolic and inflammatory blood biomarkers underscore oral health as a modifiable factor in CVD prevention. These findings highlight the clinical value of integrating multidimensional oral health assessment into cardiovascular risk stratification, especially in females and adults under the age of 65. Public health strategies aimed at preserving natural dentition, restoring masticatory function, and managing oral inflammation may serve as valuable components of broader cardiovascular risk reduction initiatives.

## Non-standard Abbreviations and Acronyms

CMEC: China Multi-Ethnic Cohort
FTU: Functional Tooth Units
NHANES III: Third National Health and Nutrition Examination Survey
OFI-8: Oral Frailty Index-8

## Acknowledgments

The authors would like to thank the individuals who participated in the CMEC cohort.

## Sources of Funding

This work was supported by the National Key Research and Development Program of China (Grant No. 2023YFC3605600) and the Research and Development Program, West China Hospital of Stomatology Sichuan University (Grant No. LCYJ-ZD-202301).

## Disclosures

The authors declare that they have no relevant disclosures.

## Notes

### Competing Interest Statement

The authors have declared no competing interest.

### Clinical Trial

ChiCTR2400082963

### Author Declarations

This study adhered to the Declaration of Helsinki, and all participants signed informed consent forms prior to data collection. Ethical approval was obtained from the Medical Ethics Review Committee of Sichuan University (WCHSIRB-D-2023-225-R1).

## References

1. Khan SS, Greenland P. Comprehensive Cardiovascular Health Promotion for Successful Prevention of Cardiovascular Disease. JAMA. 2020;324:2036–2037. doi: 10.1001/jama.2020.18731

2. Acquah I, Hagan K, Javed Z, Taha MB, Valero-Elizondo J, Nwana N, Yahya T, Sharma G, Gulati M, Hammoud A, et al. Social Determinants of Cardiovascular Risk, Subclinical Cardiovascular Disease, and Cardiovascular Events. J Am Heart Assoc. 2023;12:e025581. doi: 10.1161/JAHA.122.025581

3. Park SY, Kim SH, Kang SH, Yoon CH, Lee HJ, Yun PY, Youn TJ, Chae IH. Improved oral hygiene care attenuates the cardiovascular risk of oral health disease: a population-based study from Korea. Eur Heart J. 2019;40:1138–1145. doi: 10.1093/eurheartj/ehy836

4. Masi S, D’Aiuto F, Deanfield J. Cardiovascular prevention starts from your mouth. Eur Heart J. 2019;40:1146–1148. doi: 10.1093/eurheartj/ehz060

5. Piepoli MF, Hoes AW, Agewall S, Albus C, Brotons C, Catapano AL, Cooney MT, Corra U, Cosyns B, Deaton C, et al. 2016 European Guidelines on cardiovascular disease prevention in clinical practice: The Sixth Joint Task Force of the European Society of Cardiology and Other Societies on Cardiovascular Disease Prevention in Clinical Practice (constituted by representatives of 10 societies and by invited experts)Developed with the special contribution of the European Association for Cardiovascular Prevention & Rehabilitation (EACPR). Eur Heart J. 2016;37:2315–2381. doi: 10.1093/eurheartj/ehw106

6. Sanz M, Marco Del Castillo A, Jepsen S, Gonzalez-Juanatey JR, D’Aiuto F, Bouchard P, Chapple I, Dietrich T, Gotsman I, Graziani F, et al. Periodontitis and cardiovascular diseases: Consensus report. J Clin Periodontol. 2020;47:268–288. doi: 10.1111/jcpe.13189

7. Yu L, Hong Y, Maishi N, Matsuda AY, Hida Y, Hasebe A, Kitagawa Y, Hida K. Oral bacterium Streptococcus mutans promotes tumor metastasis through thrombosis formation. Cancer Sci. 2024;115:648–659. doi: 10.1111/cas.16010

8. Dai J, Li A, Liu Y, Chen Y, Tjakkes GE, Visser A, Xu S. Denture wearing status, cardiovascular health profiles, and mortality in edentulous patients: A prospective study with a 27-year follow-up. J Dent. 2022;126:104287. doi: 10.1016/j.jdent.2022.104287

9. Range H, Perier MC, Boillot A, Offredo L, Lisan Q, Guibout C, Thomas F, Danchin N, Boutouyrie P, Jouven X, et al. Chewing capacity and ideal cardiovascular health in adulthood: A cross-sectional analysis of a population-based cohort study. Clin Nutr. 2020;39:1440–1446. doi: 10.1016/j.clnu.2019.05.029

10. Zhao X, Hong F, Yin J, Tang W, Zhang G, Liang X, Li J, Cui C, Li X, China Multi-Ethnic Cohort collaborative g. Cohort Profile: the China Multi-Ethnic Cohort (CMEC) study. Int J Epidemiol. 2021;50:721–721l. doi: 10.1093/ije/dyaa185

11. D’Agostino RB, Sr., Vasan RS, Pencina MJ, Wolf PA, Cobain M, Massaro JM, Kannel WB. General cardiovascular risk profile for use in primary care: the Framingham Heart Study. Circulation. 2008;117:743–753. doi: 10.1161/CIRCULATIONAHA.107.699579

12. Suzuki H, Kanazawa M, Komagamine Y, Iwaki M, Jo A, Amagai N, Minakuchi S. The effect of new complete denture fabrication and simplified dietary advice on nutrient intake and masticatory function of edentulous elderly: A randomized-controlled trial. Clin Nutr. 2018;37:1441–1447. doi: 10.1016/j.clnu.2017.07.022

13. Xiang Y, Wang G, Cheng Y, Zhang Y, Zhao J, Dong B, Cai H, Cheng L, Hu T. Oral-Sarcopenia nexus in health disparities: chewing and blood mediated pathways in a Chinese aging cohort. BMC Oral Health. 2025;25:1710. doi: 10.1186/s12903-025-07059-y

14. Du M, Deng K, Yin J, Wu C, Hu S, Guo L, Luo Z, Tonetti M, Tjakkes GE, Visser A, et al. Association Between Chewing Capacity and Mortality Risk: The Role of Diet and Ageing. J Clin Periodontol. 2025;52:695–706. doi: 10.1111/jcpe.14122

15. Reue K, Wiese CB. Illuminating the Mechanisms Underlying Sex Differences in Cardiovascular Disease. Circ Res. 2022;130:1747–1762. doi: 10.1161/CIRCRESAHA.122.320259

16. Jacobs DR, Jr., Woo JG, Sinaiko AR, Daniels SR, Ikonen J, Juonala M, Kartiosuo N, Lehtimaki T, Magnussen CG, Viikari JSA, et al. Childhood Cardiovascular Risk Factors and Adult Cardiovascular Events. N Engl J Med. 2022;386:1877–1888. doi: 10.1056/NEJMoa2109191

17. Suri S, Topiwala A, Chappell MA, Okell TW, Zsoldos E, Singh-Manoux A, Kivimaki M, Mackay CE, Ebmeier KP. Association of Midlife Cardiovascular Risk Profiles With Cerebral Perfusion at Older Ages. JAMA Netw Open. 2019;2:e195776. doi: 10.1001/jamanetworkopen.2019.5776

18. Duan S, Tang R, Zhang C, Su Q, Yang H, Cai H, Hu T. The correlation of region-specific lifestyle and subjective perception of oral health with oral health-related quality of life among Tibetan early adolescents in Ganzi: a cross-sectional study. PeerJ. 2025;13:e18842. doi: 10.7717/peerj.18842

19. Demmer RT, Breskin A, Rosenbaum M, Zuk A, LeDuc C, Leibel R, Paster B, Desvarieux M, Jacobs DR, Jr., Papapanou PN. The subgingival microbiome, systemic inflammation and insulin resistance: The Oral Infections, Glucose Intolerance and Insulin Resistance Study. J Clin Periodontol. 2017;44:255–265. doi: 10.1111/jcpe.12664

20. Petrie JR, Guzik TJ, Touyz RM. Diabetes, Hypertension, and Cardiovascular Disease: Clinical Insights and Vascular Mechanisms. Can J Cardiol. 2018;34:575–584. doi: 10.1016/j.cjca.2017.12.005

21. Stark K, Massberg S. Interplay between inflammation and thrombosis in cardiovascular pathology. Nat Rev Cardiol. 2021;18:666–682. doi: 10.1038/s41569-021-00552-1

